# Antecedent Metabolic Health and Metformin (ANTHEM) Aging study: Rationale and study design for a randomized controlled trial

**DOI:** 10.1101/2021.10.20.21265196

**Authors:** Santosh Kumari, Matthew Bubak, Hayden M. Schoenberg, Arik Davidyan, Christian J. Elliehausen, Katrin G. Kuhn, Timothy M. VanWagoner, Rowan Karaman, Robert Hal Scofield, Benjamin F. Miller, Adam R. Konopka

**Author notes:** **Corresponding Author: Adam R. Konopka, PhD**, Assistant Professor, Division of Geriatrics and Gerontology, Department of Medicine, University of Wisconsin-Madison, Geriatric Research Education and Clinical Center, William S. Middleton Memorial Veterans Hospital. denotes equal contribution (Santosh Kumari, Matthew Bubak, Benjamin F. Miller, Adam R. Konopka).

## Abstract

The antidiabetic medication metformin has been proposed to be the first drug tested to target aging and extend healthspan in humans. While there is extensive epidemiological support for the health benefits of metformin in patient populations, it is not clear if these protective effects apply to those free of age-related disease. Our previous data in older adults without diabetes suggest a dichotomous change in insulin sensitivity and skeletal muscle mitochondrial adaptations after metformin treatment when co-prescribed with exercise. Those who entered the study as insulin sensitive had no change to detrimental effects while those who were insulin resistant had positive changes. The objective of this clinical trial is to determine if 1) antecedent metabolic health and 2) skeletal muscle mitochondrial remodeling and function mediate the positive or detrimental effects of metformin monotherapy, independent of exercise, on the metabolism and biology of aging. In a randomized, double blind clinical trial, adults free of chronic disease (n=148, 40-75 years old) are stratified as either insulin sensitive or insulin resistant based on HOMA-IR (≤2.2 or ≥2.5) and take 1500 mg/day of metformin or placebo for 12 weeks. Hyperinsulinemic-euglycemic clamps and skeletal muscle biopsies are performed before and after 12 weeks to assess primary outcomes of peripheral insulin sensitivity and mitochondrial remodeling and function. Findings from this trial will identify clinical characteristics and cellular mechanisms involved in modulating the effectiveness of metformin treatment to target aging that could inform larger phase 3 clinical trials aimed at testing aging as an indication for metformin.

## Introduction and Rationale

The dramatic demographic shift due to world-wide population aging is one of the most critical societal challenges of our time^1^. Within the next 30 years, the number of people over the age of 65 is estimated to reach 2 billion^2^. Increased longevity is accompanied by a parallel increase in the incidence of chronic diseases^2^. Age is a primary risk factor for most chronic conditions, including type 2 diabetes (T2D), cardiovascular disease, arthritis, sarcopenia, dementia, and cancer^3,4^. Additionally, ∼63% of the population ≥65 years old has 2 or more age-related chronic conditions which is nearly double that of people aged 45-64 years old^5^. A recent framework has divided lifespan into a time of life free of disease (healthspan) followed by the accumulation of one or more overt age-associated diseases and disabilities^4^. There is an urgent need to develop strategies that can simultaneously decrease the risk of age-related co-morbidities. Such strategies that delay the onset of age-related chronic conditions to improve healthspan must start before the onset of age-related disease^6^. It is now critical to test new or repurposed existing drugs for healthspan extension in people who are currently free of chronic disease.

Metformin is a candidate to be the first pharmaceutical treatment to slow aging and extend healthspan^7^. Metformin is the most widely prescribed oral anti-hyperglycemic medication and has 60 years of safety documentation, low cost and is associated with decreased risk of mortality and diseases such as T2D, cardiovascular disease (CVD), dementia and cancer^8–12^. While these findings provide rationale and fuel the excitement for metformin to treat aging, the completed studies were conducted in subjects with T2D and none were in relatively healthy subjects, absent of disease. Thus, the efficacy of metformin to improve outcomes associated with the aging process in human subjects free of chronic disease remains incompletely understood.

While the pleiotropic effects of metformin treatment could potentially be advantageous for targeting the basic biology of aging, the health and lifespan extending effects of metformin are equivocal across multiple models, including Caenorhabditis elegans (*C*.*elegans*), mice, rats, and humans. Metformin increases mean lifespan when started in young *C*.*elegans*^16,17^, yet the opposite is true in aged *C*.*elegans* where metformin decreases lifespan^18^. However, insulin resistant *daf-2(e1370)* mutant *C*.*elegans* were impervious to the decrease in lifespan by metformin^18^. Dietary metformin (1000ppm) increased lifespan and indices of healthspan in male C57BL6 mice^19^ but not in male and female UMHET3 mice^20^. Metformin can even shorten lifespan in C57BL6 mice when used at higher dietary doses (10,000ppm)^19^. Collectively, these studies in model systems highlight that the positive effects of metformin are not universal and it remains unclear if metformin will be an effective treatment to improve aging processes in people free of chronic disease.

The landmark Diabetes Prevention Program trial examined the progression of subjects with pre-diabetes to T2D^21^. Over the three years of study, the metformin treatment group (1700mg/day; n=1073) had significantly less progression to T2D compared to controls. However, the effect of metformin was minimal in those with a lower BMI and lower fasting glucose and tended to be less effective in older versus younger adults^21,22^. These findings are in line with our recently completed randomized, double-blinded clinical trial investigating the impact of metformin on the healthspan-extending effects of exercise^24^. Sedentary older subjects (n=53, 62±2 yrs) completed 12-weeks of aerobic exercise training while taking either placebo or metformin (1500-2000mg/day). On average, metformin prevented the expected exercise-induced increase in whole-body insulin sensitivity and mitochondrial complex I linked respiration. However, there was a wide range of inter-subject heterogeneity specific to the metformin group, but not the placebo group, where some subjects demonstrated the expected exercise-induced improvement in insulin sensitivity while others had no improvement or a decrement in insulin sensitivity^24^. Retrospectively, we rank sorted several characteristics of subjects in the metformin group to determine if any baseline characteristics were associated with the change in insulin sensitivity after exercise training and metformin treatment (**Figure 1**). There were significant differences in the change in insulin sensitivity associated with baseline mitochondrial complex I respiration, insulin sensitivity and HOMA-IR, when subjects were ranked and divided into two groups representing the upper 50% (high) and lower 50% (low). In other words, negative responders were subjects that began the study with greater mitochondrial complex I respiration and insulin sensitivity and lower HOMA-IR. We interpreted these secondary analyses to suggest that subjects that entered the study in a metabolically healthy state had detrimental changes in insulin sensitivity when taking metformin with exercise, while those that were relatively less metabolically healthy had positive changes. We do not yet know if these detrimental or positive responses happen with metformin monotherapy in the absence of exercise training. Therefore, if metformin is to be recommended to slow the onset of age-related disease, we need to further understand the characteristics of subjects that do or do not benefit from metformin treatment.

**Figure 1.**
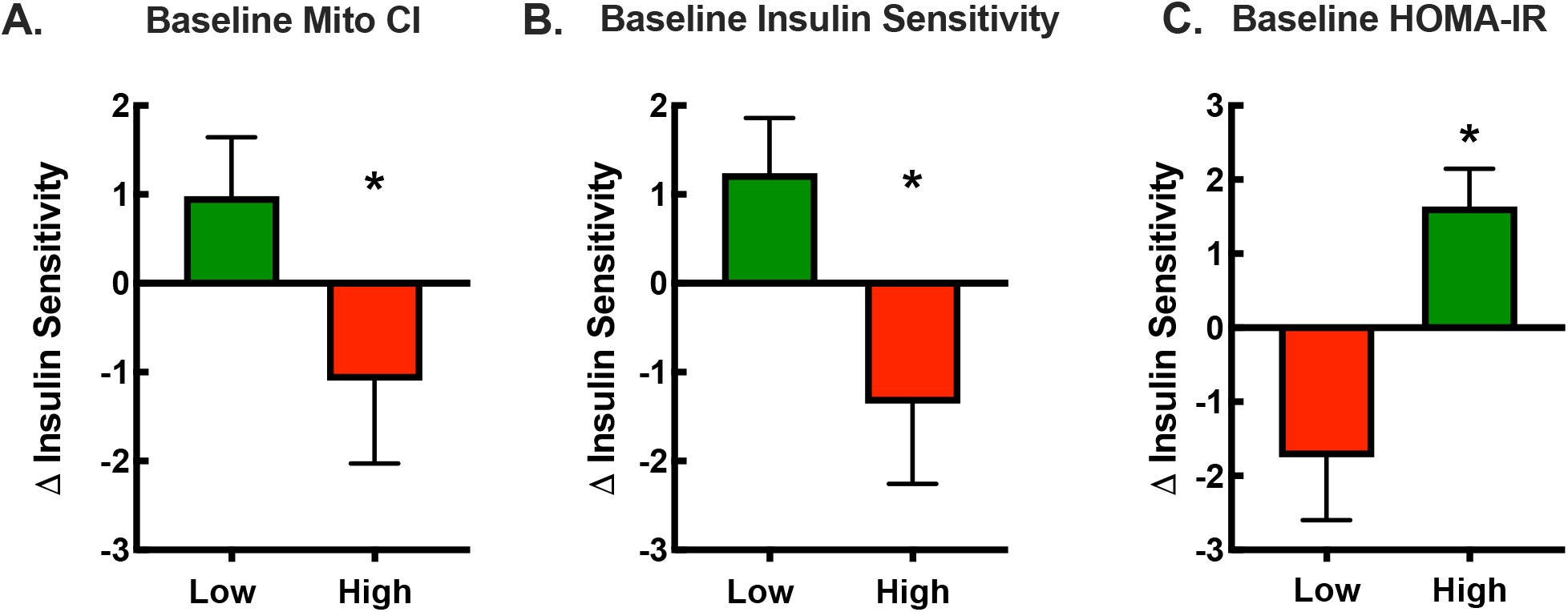
Retrospective analysis of baseline subject characteristics on the change in insulin sensitivity after metformin plus exercise. Subjects (n=25-27) from our previous study^26^ were divided into two groups representing the upper 50% (high) and lower 50% (low) for baseline **A)** skeletal muscle mitochondrial complex I-linked (CI) respiration, **B)** insulin sensitivity and **C)** HOMA-IR. The negative responders, noted in the red bars, were the subjects that began the study with greater mitochondrial CI-linked respiration, higher insulin sensitivity, and lower insulin resistance. The positive responders, noted in green bars, were the subjects that began the study with lower mitochondrial CI-linked respiration, lower insulin sensitivity and greater insulin resistance. These data suggest those who were metabolically healthy had a negative response while those who were relatively less healthy had a positive response to metformin adjuvant to exercise. As previously described^26^, we performed skeletal muscle biopsies to assess skeletal muscle mitochondrial respiration in permeabilized fibers, a 75g oral glucose tolerance test to determine insulin sensitivity via the Matsuda Index and fasting insulin and glucose values to determine insulin resistance via HOMA-IR.

Skeletal muscle is the largest organ in the body and is critical for maintaining physical function, energy balance, and metabolic health across the lifespan. With increasing age, there is an association between skeletal muscle mitochondrial respiration and walking speed^25^, aerobic capacity^26,27^, muscle strength^27^ and insulin sensitivity^26^. However, as previously reviewed^6,28^, studies evaluating the effects of metformin on skeletal muscle mitochondria are limited with nearly an equal number of studies supporting or refuting the idea that metformin inhibits complex I activity. Studies that suggest an inhibitory effect of metformin on skeletal muscle mitochondria have used supra-pharmacological doses^29–31^. Our data were the first to show that a clinically relevant dose of metformin prevented the expected increase in complex I-linked mitochondrial respiration after aerobic exercise training^24^. Whether metformin acts directly on the mitochondria or indirectly through mitochondrial protein remodeling remain unclear. We have previously shown that adding metformin to rapamycin in young UMHET3 mice decreases protein synthesis rates of complexes within the electron transport system, including proteins belonging to complex I^32^. Therefore, it remains to be reconciled if complex I protein remodeling is a mechanism by which metformin monotherapy modulates skeletal muscle respiration and hydrogen peroxide (H_2_O_2_) emissions.

This clinical trial will address two unresolved questions: 1) does antecedent metabolic health influence healthspan-related responses to metformin monotherapy, and 2) does metformin treatment lead to mitochondrial protein remodeling in skeletal muscle that changes mitochondrial function. We hypothesize that metformin treatment will improve insulin sensitivity and glucoregulation in insulin resistant individuals, but will decrease or not improve insulin sensitivity and glucoregulation in insulin sensitive subjects. We also hypothesize that metformin treatment will remodel skeletal muscle mitochondria in a way that improves mitochondrial function in subjects that are insulin resistant but decreases mitochondrial function in subjects that are insulin sensitive. We will also explore if proposed biomarkers of aging^33^ predict or correlate with changes in insulin sensitivity and mitochondrial metabolism in disease-free individuals. This is the first human clinical trial aimed at identifying the clinical and cellular characteristics that modulate the effectiveness of metformin on the metabolism and biology of aging in individuals free of disease.

### Study Overview

We have started a 12-week randomized, double-blind, placebo-controlled clinical trial at two sites; 1) Oklahoma Medical Research Foundation/University of Oklahoma Health Sciences Center (collectively referred to as OKC) led by BFM and 2) the University of Wisconsin-Madison (UWM) led by ARK. The ongoing trial is registered at clinicaltrials.gov (NCT04264897). We are recruiting 148 participants, including men and women of all races and ethnic backgrounds between the ages of 40 and 75 who are not being treated with glucose-lowering drugs nor have overt chronic diseases (**Table 1**). Participants are eligible if they have impaired fasting glucose, obesity, hypertension, hyperlipidemia, or are physically inactive or on most medications (e.g., statins) treating risk factors for overt cardiometabolic disease. Fifty percent of subjects will be recruited at each site (OKC and UWM), where half of the subjects are stratified as insulin sensitive (n=74) and half as insulin resistant (n=74) based on baseline HOMA-IR (≤2.2 or ≥2.5). As illustrated in **Figure 2**, eligible particpants will complete a DEXA scan to assess body composition, continuous glucose monitor wear period to evaluate ambulant glucose behavior, a skeletal muscle biopsy to determine mitochondrial respiration, hydrogen peroxide emissions and protein remodeling and a 3-hr hyperinsulinemic-euglycemic clamp to measure peripheral insulin sensitivity. Details of the study design, particpants, and methodology can be found in the **online supplement**.

**Table 1:**
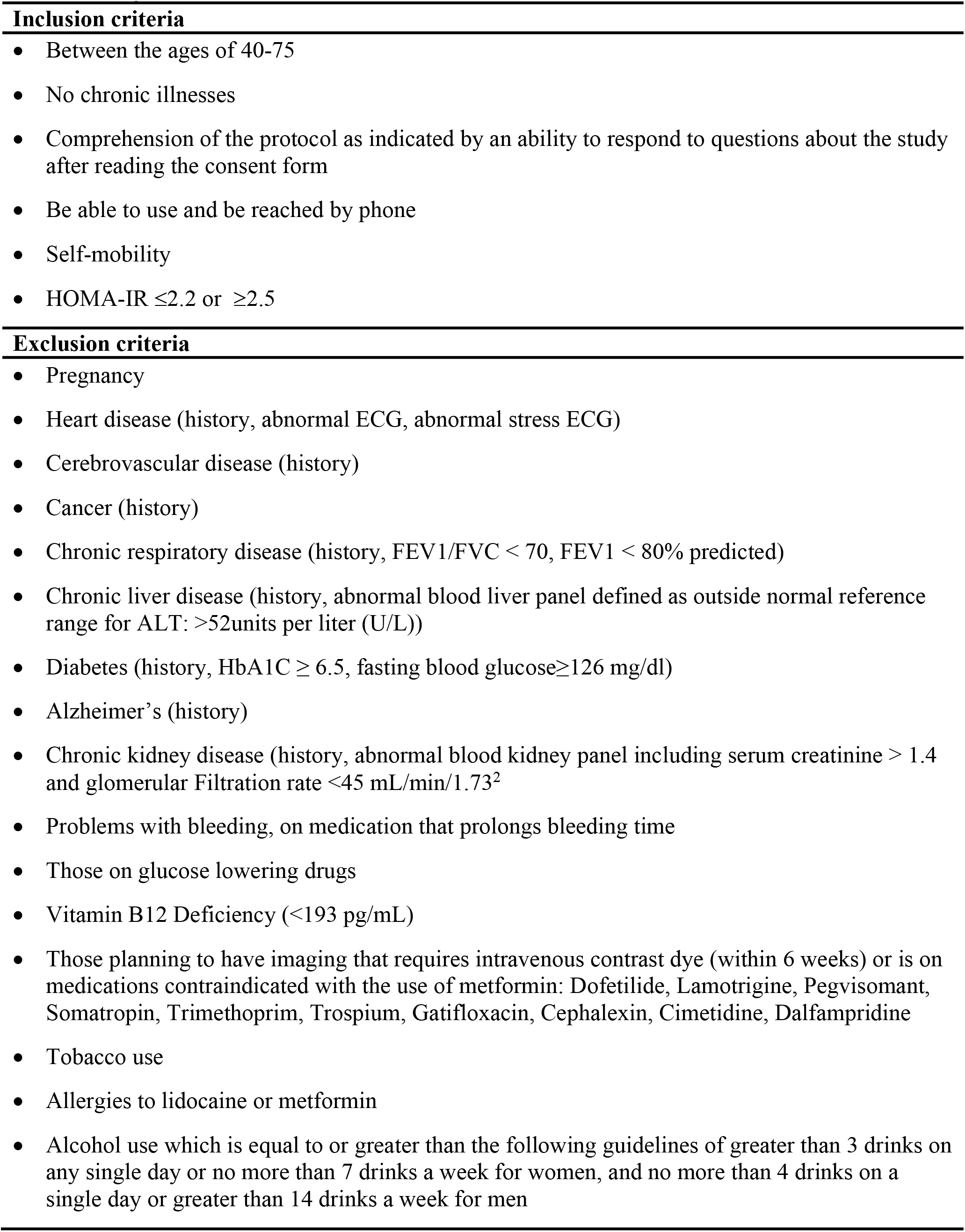
Study inclusion and exclusion criteria

**Figure 2.**
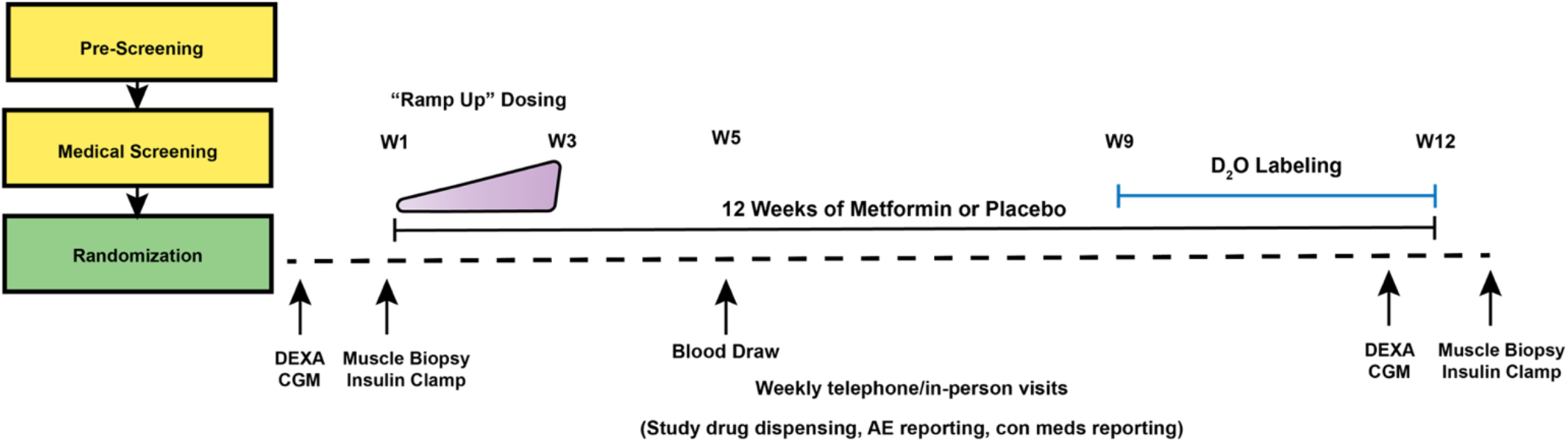
Study Design Schematic. To understand the translational potential of metformin to slow aging, this ongoing 12-week, double-blinded, randomized control trial aims to determine how antecedent metabolic health (insulin resistant vs. insulin sensitive) and the relationship between skeletal muscle mitochondrial remodeling and function impact the potential healthspan extending effects of metformin. After screening, eligible participants will be randomized to 12 weeks of placebo or metformin (1500 mg/day) by the study statistician. Pre and post-intervention assessments include dual x-ray absorptiometry (DEXA) to assess body composition, continuous glucose monitoring (CGM) to determine glucoregulation, proposed blood-based biomarkers of aging, hyperinsulinemic-euglycemic clamp to measure peripheral insulin sensitivity and muscle biopsy samples to evaluate mitochondrial function by high-resolution fluorespirometry. We will also use deuterium oxide (D_2_O) stable isotope labeling during the last 4 weeks of the intervention to determine individual and complex-specific mitochondrial remodeling from post muscle biopsy samples. Abbreviations: DEXA, Dual X-ray Absorptiometry; CGM, Continuous glucose monitor; W, Week; Con Meds, concomitant medications.

## Conclusion

There is an urgent need to develop new therapeutic options to prevent or delay the deleterious changes of aging that contribute to morbidity among older adults. As we have illustrated, there are contradictory findings about chronic metformin treatment, and the effects of metformin treatment in those free of disease are largely unknown. Although there are studies using metformin to delay morbidity in rodent models, there is a noted lack of investigations examining metformin use in a middle-to-older-aged human population absent of metabolic disease. Importantly, within a population of disease-free individuals there is a range of metabolic health and insulin sensitivity, which makes some more at risk for chronic age-related disease than others. With the geroscience goal of preventing or delaying morbidity and mortality, it is implied that treatment should begin before the accumulation of age-related morbidities. Therefore, our study will close the knowledge gap in understanding how metformin may affect aging related processes in subjects free of disease and identify the mechanisms through which metformin has potentially varying effects on aging-related outcomes. By the conclusion of this study, we will have provided insight into which subjects could benefit from metformin treatment and thereby inform the design of a larger clinical trial aimed at using metformin to target aging.

## Data Availability

All data produced in the present study will be available upon reasonable request to the authors

## Acknowledgements

This study is ongoing and we would like to thank our participants for their time and dedication. We appreciate the efforts of Jaime Laurin and Katlyn Beecken at the initiation of the study and Dr. Julie Stoner (deceased) for assistance with creating the study protocol and statistical design. We also like to thank Gina Herbert at OKC and Sara Decker, Tamara Kempken Mehring, Tammy Kiger, and Susan Johnston within the Clinical Research Unit and the Pharmaceutical Research Center at UWM for their detailed implementation and execution of the study protocol. We would also express our gratitude to Samantha Lebsock at Belmar Pharmacy for leading study drug compounding and allocation. We apologize that we could not cite many studies using metformin due to reference limitations.

ARK is the Co-I for the project and PI for the UWM site. BFM is the PI for the project and OKC site. ARK and BFM are responsible for study design, study execution, data collection, data analysis and data interpretation. SK, MB, ED, CJE, TMV are responsible for data collection and analysis. KGK is the biostatistician and responsnible for randomization and data analysis. RK and RHS are study physicians and provide medical oversight.

## Supplemental Information

### Methods

The University of Oklahoma Health Sciences Center serves as the single IRB of record and approved all study procedures. The study is monitored by an external Data Safety Monitoring Board that was appointed by the National Institute on Aging. To ensure fidelity between OKC and UWM study sites, we have performed multiple site visits, exchanged reagents, and have monthly study-specific virtual meetings to address recruitment, enrollment, study protocol deviations, amendments and troubleshooting.

### Study Population

We are recruiting 148 participants (40-75yrs old) who are either insulin sensitive or insulin resistant defined by a HOMA-IR of ≤2.2 or ≥2.5. Epidemiological studies using diverse subjects without diabetes have defined insulin resistance using HOMA-IR values of 1.5-3.0^1^. Therefore, we chose the midpoint of these HOMA-IR values ≤2.2 to determine insulin sensitivity and ≥2.5 to determine insulin resistance. Since some studies have found that HOMA-IR values of ≤2.0 define insulin resistance, an alternative exploratory approach will be to assess how alternative thresholds for insulin resistance based on HOMA-IR impact metformin treatment on primary and secondary outcomes. Importantly, HOMA-IR has been successfully used to stratify participants when confirmed using a hyperinsulinemic-euglycemic clamp^2^ as performed in the current study.

### Pre-Screening

Individuals expressing interest in participating in the study are contacted by the study team to determine potential eligibility using general inclusion/exclusion criteria questions and self-reported medical history. If subjects are eligible via telephone screening, they are scheduled for health and medical screening and are sent an electronic copy of the informed consent document to review.

### Informed Consent, Health and Medical Screening

After documentation of informed consent, we query subjects for their health and physical activity history, including any medical conditions, recent illnesses, COVID-19 symptoms, and medications. Height and weight, resting blood pressure, and heart rate are also collected. Women of childbearing capacity complete a urine pregnancy test. Participants complete medical screening, which consists of a fasting blood draw for HbA1c, glucose and insulin, complete metabolic panel (CMP), Vitamin B12, resting 12-lead ECG, and a pulmonary function test. Supervising study physicians at each site (RK and RHS) will review all information and determine subject eligibility, as defined in **Table 1**. We use the objective distinction of chronic disease versus risk factors for chronic disease as defined by both the CDC and WHO. We will screen subjects to eliminate those with a chronic disease (e.g. cardiovascular disease, type 2 diabetes), but will include people with the following risk factors for cardiometabolic disease: family history, physical inactivity, obesity, hyperlipidemia, hypertension, and impaired fasting glucose (>126 mg/dL). **Table 1** details the screening procedures and objective criteria overseen by the study physician for determining if subjects are free of chronic disease. Table 1 also includes a list of drugs or drug classes that have been reported to be contraindicated with metformin, safety concerns with study procedures, or impact the primary outcomes. We are including subjects that use commonly consumed medications to control cholesterol and blood pressure. For example, we have previously found that ∼40% of subjects are on statins but they have not responded differently to metformin than non-statin users^3,4^.

### Randomization

The study biostatistician (KGK) randomizes eligible, consenting participants to metformin and placebo (1:1). The randomization sequence is stratified by study center (OKC or UWM) and baseline insulin resistance status (HOMA-IR ≤2.2 versus ≥2.5) and created using randomly chosen block sizes of four or six (STATA 16). Only the biostatistician and pharmacy have access to the randomization and stratification scheme. Therefore, all members of the investigative team will remain blinded throughout data collection and analysis.

### Body Composition and Continuous Glucose Monitoring

Eligible subjects complete body composition assessments via dual x-ray absorptiometry (DXA; Lunar iDXA, GE Electric, Boston, MA). After the DXA scan, all subjects have the choice to opt in to wearing the continuous glucose monitor (CGM; Dexcom G6 Pro) to evaluate glucose behavior and variability during 7-10 days of free-living conditions that complements our gold-standard laboratory assessments of glucose control and insulin sensitivity. The CGM provides a high-resolution assessment of glucose behavior by measuring interstitial glucose values every 5 minutes (288 data points per 24 hours). The Dexcom G6 CGM has a mean absolute relative difference (MARD) to clinical reference values of 9% across a 10 day wear period^5^. Glucose variability, even in the absence of sustained hyperglycemia, has been shown to have deleterious effects, such as increased oxidative stress, that are associated with aging and chronic diseases^6^. Participants track their physical activity using a 7-day physical activity recall, which for a subset of participants overlaps the CGM wear period. Participants repeat the DXA scan and CGM wear period during the last 7-10 days of study drug leading up to the post-intervention skeletal muscle biopsy and hyperinsulinemic-euglycemic clamp.

### Skeletal Muscle Biopsy and Hyperinsulinemic-Euglycemic Clamp

All muscle biopsies and hyperinsulinemic-euglycemic clamps are completed in the Clinical Research Units (CRU) at the Oklahoma Clinical and Translational Science Institute (OCTSI) at OKC and the Institute of Clinical and Translational Research (ICTR) at UWM. A skeletal muscle biopsy and hyperinsulinemic euglycemic clamp are performed once before and repeated once after the 12-week intervention. The post muscle biopsy and insulin clamp are completed approximately 36 hours after the last study drug administration to understand the long-term effects of metformin and not the acute response.

Subjects are asked to refrain from exercise, alcohol, and aspirin for 24 hours prior to the muscle biopsy and insulin clamp. The night before, participants consume a standardized study meal provided by the study team. The meals (∼750 kcal) before and after the intervention are matched for macronutrient composition (40% carbohydrates) to minimize inter and intra-subject variability. Subjects will arrive to the CRU the next morning (∼0700) after an overnight fast where body weight and vitals (temperature, heart rate, and blood pressure) are recorded. After resting for ∼30 minutes, muscle samples (100-300mg) are obtained from the *vastus lateralis* after administration of local anesthetic (1-2% lidocaine without epinephrine) using a 5mm UCH needle (Millennium Surgical) with manual suction^3,7^. Pressure is held for 10 minutes before the incision is closed with steri-strips and a pressure bandage is applied and worn the remainder of the day. Muscle samples are placed on a culture dish on ice and cleared of visible adipose and connective tissue. Samples are subsequently incubated in ice-cold Buffer Z for fresh tissue mitochondrial function analyses or snap frozen in liquid nitrogen and stored at -80°C for subsequent quantitative and kinetic proteomics.

Approximately 30-45 minutes after the muscle biopsy is performed, an intravenous catheter is placed in a heated hand vein for repeated arterialized-venous blood sampling and in the contralateral forearm for infusion of insulin and dextrose. Blood is obtained before the start of insulin and dextrose infusions for the measurement of glucose, glucoregulatory hormones and proposed biomarkers of aging. Next, peripheral insulin sensitivity is measured during a 180 min hyperinsulinemic-euglycemic clamp similar to previously described procedures^8–10^. Insulin (2.3 mU·kgFFM^−1^·min^−1^) is infused to mimic post-prandial insulin values and dextrose (25%) is infused at a variable rate to maintain euglycemia, defined here as a target range of 85-95 mg/dL. Our study will measure insulin sensitivity based on the glucose infusion rate to maintain euglycemia during the last 30 minutes as well as exploratory analysis of total or incremental area under the curve (AUC_180_, AUC_120_, AUC_60_).

### Study Drug

Belmar Pharmacy has compounded both metformin (NDC: 23155-102-01) and placebo (PROSOLV^®^ SMCC, Sodium Starch Glycolate, Magnesium Stearate), which is then shipped to each study site pharmacy to dispense to enrolled subjects. The study biostatistician (KK) communicates randomization directly with Belmar Pharmacy to maintain study blind. Metformin and placebo are provided in tablet form and dosed as 500 mg tablets. Study drugs are equal in size and shape, with no distinguishing marks. Study drug is administered using a ramped protocol, starting at 500 mg per day in week 1, increasing to 1000 mg per day in week 2, and then to 1500 mg per day in week 3, as tolerated. The dose is maintained at 1500 mg/day for the remaining 9 weeks. Two 500 mg tablets are taken in the morning, and one 500 mg tablet taken in the evening. After 5 weeks of study drug, participants will complete an additional fasting blood draw for CMP and Vitamin B_12_ to ensure maintenance of normal reference values. If a subject experiences gastrointestinal discomfort or other common side effects associated with metformin while taking 1500 mg per day, the dose is reduced to 1000 mg per day. The minimal dose required to remain enrolled in the study is 1000 mg/day. We have previously found that subjects in both placebo and metformin report symptoms commonly associated with metformin side effects and therefore reporting these symptoms do not jeopardize the study blind^3,4^.

### Adverse Reaction Monitoring

Although metformin is commonly used to treat diabetes and is safe, there are some notable side effects. Lactic acidosis is the most serious known adverse event and occurs most frequently in patients with impaired liver, kidney or heart function. Those with liver, kidney or heart conditions will be screened out from our study and minimize the risk of lactic acidosis. The most common and less serious side effects of metformin include gastrointestinal distress including loss of appetite, nausea, vomiting, diarrhea, flatulence, or stomach discomfort. There is also a risk of Vitamin B12 deficiency. Participants will be screened for Vitamin B12 levels at baseline and after 5 weeks of study drug.

### Deuterium Oxide Labeling and Mitochondrial Protein Turnover

To understand long-term mitochondrial protein remodeling, we use deuterium oxide (D_2_O) labeling to measure protein turnover as we have previously published^3,7,11,12^. The principle of the method is the same as other approaches that rely on precursor and product labeling over a period of time to measure a rate of synthesis. In this case, provision of D_2_O daily can maintain a steady state body water enrichment where the heavy hydrogens equilibrate with nonessential amino acids through pathways of intermediary metabolism. In our study, the subjects consume 70% D_2_O (Millipore Sigma Cat# 756822) 3 × 50 ml/day for one week followed by 2 × 50 ml/day for 3 weeks to maintain a body water enrichment of 1-2%. At the completion of the labeling period, the precursor enrichment is determined from body water enrichment of plasma from each subject, while the product enrichment is determined from proteins of the skeletal muscle biopsy. For this study, we will be using a targeted proteomics approach that allows assessment of the change in concentration and synthesis rates of individual mitochondrial proteins. By measuring both the change in protein concentrations of individual mitochondrial proteins and the rates of synthesis of those individual proteins, we can also calculate protein breakdown. To accomplish these measurements, approximately 20 mg of the muscle biopsy will be analyzed using targeted proteomics of mitochondrial proteins by LC-MS/MS (ThermoScientific Q-Exactive Plus). These analyses will allow us to determine differences in mitochondrial remodeling in skeletal muscle mitochondria with metformin treatment.

### Skeletal Muscle Mitochondrial Bioenergetics

We prepare permeabilized fibers for the assessment of skeletal muscle mitochondrial bioenergetics as previously performed^3,8,11,13^. Since complex I activity is one purported mechanism by which metformin regulates the biology and metabolism of aging, our primary objective is to evaluate complex I-linked submaximal and maximal mitochondrial respiration at both study sites using a similar ADP titration as described previously^3,11,13^. At OKC, there will be an additional protocol using complex I and II substrates (pyruvate-glutamate-malate and succinate) to explore maximal mitochondrial hydrogen peroxide (H_2_O_2_) emissions. We will determine the sensitivity of mitochondria to ADP to stimulate 50% maximal respiration and to suppress maximal H_2_O_2_ emissions by 50% from Michaelis-Menten kinetics and one-phase exponential decay analysis to calculate the apparent K_m_ and half maximal inhibitory concentration (IC_50_) for ADP^14^. At UWM, we will use a creatine kinase energetic clamp technique described in detail by Fisher-Wellman et al.^15^ to model in vivo demands for ATP. Most approaches that assess maximal respiration through an ADP bolus create an ATP/ADP ratio that is not physiologically relevant. The creatine kinase clamp exploits the enzymatic activity of creatine kinase to titrate the extra-mitochondrial ATP/ADP ratio to evaluate whether metformin is changing the mitochondrial respiratory control and conductance across a range of physiological ATP free energy states.

### Oxidative Stress Measures

Metformin can alter redox status in the liver^16^, while the impact on skeletal muscle is less clear. To determine the redox status of skeletal muscle, we will assess the redox pairs GSH/GSSG, NADPH/NADP+, NADH/NAD+ by ion pairing reverse-phase HPLC quantified using electrochemical, fluorescence, or UV/VIS detection^17^. For a sensitive measure of oxidative damage, we will assess lipid peroxidation by measuring F_2_-isoprostanes using GC/MS^17^.

### Biomarkers of Aging

IL-6, CRP, Tumor Necrosis Factor-RII, GDF15, IGF-1, Cystatin-C, B-type natriuretic peptides (NT-proBNP), and HbA1C were recently suggested as blood-based biomarkers for the use in geroscience clinical trials^18^. To determine whether the proposed biomarkers of aging predict changes in disease-free individuals by metformin treatment, we will assess these outcomes pre and post intervention. These analyses will be performed using the Bioplex 200 bead array system for xMAP in a 4-plex format in a 96 well plate. Some biomarkers will be assessed in plasma samples using analyte-specific direct ELISAs that are commercially available. These analyses will be compared to control (matched age, sex, race) plasma samples. All samples will be randomized across plates in order to control for bias. ELISAs will be read on standard plate readers and will be assessed using the manufacturer-recommended analytical parameters.

### Statistical approach

#### Sample size and power calculations

The primary outcomes from this study are insulin sensitivity and mitochondrial respiration. Based on preliminary data from a 12-week treatment period with metformin, the mean (SD) change in insulin sensitivity (post-minus pre-treatment) was 0.67 (2.86) among insulin sensitive (HOMA-IR≤2.5) and 1.36 (1.85) among insulin resistant individuals^3^. Assuming no change on average among the placebo-treated participants, a total sample size of 148 (37 per insulin sensitivity group at each site) will result in 80% power to detect an interaction between metformin treatment and baseline insulin sensitivity where the root mean square term (standard deviation) for the interaction is 0.65 or greater (an effect size of 0.25) (PASS13, NCSS, Utah, USA). This calculation is based on a 2×2 factorial ANOVA model and assumes a within-group root mean square error of 2.64, a two-sided 0.05 alpha level, and a withdraw rate of no more than 10% over the 12-week treatment period.

Based on preliminary data from a 12-week treatment period with metformin, the mean (SD) change in the complex I activity (post-minus pre-treatment; pmol•s^-1^•mg tissue^-1^) was 5.86 (16.91) among insulin resistant (HOMA-IR ≥ 2.5) and -0.28 (21.97) among insulin sensitive individuals^3^. Among participants receiving placebo, the estimates were 9.87 (11.93) and 12.01 (15.87), respectively. A total sample size of 148 (37 per insulin sensitivity group at each center) will result in 80% power to detect an interaction between metformin treatment and baseline insulin sensitivity where the root mean square term (standard deviation) for the interaction is 4.0 or greater (an effect size of 0.25) (PASS13, NCSS, Utah, USA). This calculation is based on a 2×2 factorial ANOVA model and assumes a within-group root mean square error of 16.35, a two-sided 0.05 alpha level, and a withdraw rate of no more than 10% over the 12-week treatment period. Similarly, the targeted sample size will result in 80% power to detect an interaction between metformin treatment and baseline insulin sensitivity where the root mean square term (standard deviation) for the interaction is 5.25 or greater (an effect size of 0.25) and the within-group root mean square error is 20.73 based on the ADP titration protocol with mean (SD) among metformin-treated participants of 7.65 (22.85) and -10.19 (17.40) for IR and IS participants, and among placebo-treated participants of 19.43 (23.14) and 6.68 (32.69), respectively.

#### Statistical analysis plans

The mean change in insulin sensitivity (calculated as the 12-week minus baseline measure) will be compared between groups defined by baseline insulin sensitivity (IR or IS) and metformin/placebo assignment using a 2-factor ANOVA model that includes the treatment by insulin sensitivity interaction. A significant interaction will be followed by analyses stratified by baseline insulin sensitivity. Transformations will be used as appropriate to satisfy modeling assumptions. Analyses will be based on the intention-to-treat principle. A separate per-protocol analysis, analyzing data from all participants who took at least 80% of the pills, will also be performed. Sex will be investigated as a biologic variable by including sex, and interactions with sex, in the ANOVA model. Modification by sex will be explored, however, interpreted cautiously given that the study is not powered to detect modification by sex. A similar approach will be used to explore age, BMI, and physical activity as biologic factors. A mixed effects ANOVA model will be used for the other outcome measures (utilizing day-level average measures) where the model includes fixed effects for baseline insulin sensitivity category, metformin/placebo assignment, time (baseline or 12-week), and all possible interaction effects. A random participant effect also will be included. The primary term of interest is the time by insulin sensitivity by treatment interaction (indicating that the effect of metformin on the outcome following the 12-week treatment period differs depending on baseline insulin sensitivity). The mixed effects ANOVA modeling for repeated measures described for insulin sensitivity will be used to analyze mitochondrial function. Statistical significance will be declared at p<0.05. For quantitative proteomics, we will use our previously published methods using a Benjamini-Hochberg false discovery rate correction (q = 5%)^19,20^. Heatmap and principal component analysis (PCA) plot will be constructed with ClustVis with default settings (Row scaling = unit variance scaling, PCA method = SVD with imputation, clustering distance for rows = correlation, clustering method for rows = average, tree ordering for rows = tightest cluster first).

